# Experiences of family caregivers in caring for patients with heart failure at Jakaya Kikwete Cardiac Institute, Dar es Salaam, Tanzania: a qualitative study

**DOI:** 10.1101/2023.01.12.23284350

**Authors:** Tunzo L. Mcharo, Masunga K. Iseselo, Samwel E. Kahema, Edith AM. Tarimo

## Abstract

**Background:** Heart failure (HF) continues to be a global health problem with its ramifications more pronounced in underdeveloped countries. Family members play a pivotal part in patient management which may influence the patient’s overall quality of life. Prolonged delay in attendance to health care facilities among patients indicates ineffective support from family caregivers. In the Tanzanian context, there is limited information about the experiences of family caregivers in caring for patients with HF. This study explored family caregivers’ experiences in caring for HF patients.

**Methods:** A qualitative descriptive study design was conducted at Jakaya Kikwete Cardiac Institute in Dar es Salaam, Tanzania. A purposive sampling technique was used to select the potential participants. A sample size of 10 family caregivers of patients with HF was included in the study. Thematic analysis was used to derive the main theme and sub-themes.

**Results:** Three major themes were identified: demands for supportive care, new caring role and lifestyle, and professional support in caring for patients with HF. Caregivers needed social and financial support to facilitate the caring process. Learning to provide the required care at the right time was the new role acquired by caregivers while failing to participate in social events and caregiving in an unfavourable environment were reported as challenges in caregiving. However, compliance with instruction and effective interaction among the nurses and caregivers were considered to be positive professional support.

**Conclusion:** Caregivers need social and financial support to provide effective care to their patients. Caregiving is a learning process that needs continuous educational support to adapt to the new caring roles and challenges. Nurses should conduct regular assessments to explore caregivers’ needs, challenges, and concerns and provide timely counselling that can facilitate coping.

## Introduction

Heart failure (HF) is a complex clinical illness which is brought on by a structural or functional impairment of ventricular filling or blood ejection. (1). Heart failure is a widespread problem that has serious consequences for public health. According to a Global Health Data Exchange (GHXx) report, the current global prevalence of HF is 64.34 million cases (8.52 per 1,000 inhabitants) (2). In the United States of America, HF affects 6 million adults (3,4). In the United Kingdom (UK) HF affects 900,000 annually, is predominantly a disease of older people and affects at least 5% of those aged 75 years old rising to 15% in very old people (5).

In Sub-Saharan Africa (SSA) HF is linked to high rates of recurrent hospitalization, morbidity and mortality, poor quality of life, and lost economic productivity (6). The majority of young and prosperous adults are affected with HF (6,7) with 75% of cases in 2005 being non-ischemic (7). Heart diseases, including HF, are common in Tanzania (8). Contrary to developed nations where ischemic heart disease predominates, Tanzania’s leading contributor to HF is hypertension (8). This emphasizes the importance of early detection and treatment of hypertension. Anaemia, atrial fibrillation, and a lack of education are modifiable predictors of mortality (8).

Family and friends participate in the management of patients with HF (4). Patients with HF usually depend on family members to carry out everyday tasks and manage their treatment process due to their poor physical and cognitive health (4,9). To maintain patients’ adherence and self-care behaviour, family caregivers play a crucial role in community-based HF management, which requires a significant time and energy investment (10).

Families are anticipated to play a bigger part in the healthcare system (11). Patients with HF rely on family caregivers for help with self-care behaviours and day-to-day monitoring of health status. The prolonged delay in attendance to healthcare facilities among patients indicates ineffective support from family caregivers (12). Although progress has been made in studying and documenting the experiences of family caregivers in caring for patients with HF (10), up to date, there is limited information on the experiences of the family caregiver in caring for patients with HF in the Tanzania context. High-quality care for HF patients includes effective support from family caregivers (13).

## Materials and Methods

### Study Setting

The study was conducted at Jakaya Kikwete Cardiac Institute (JKCI) in Dar es Salaam, Tanzania. JKCI is a nationally recognized teaching hospital with a focus on cardiovascular treatment, education, and research. The Institute can accommodate 120 patients, and on average, there are 700 outpatients and 100 inpatients each week with different heart conditions including HF (14). This facilitated easy access to the study participants.

### Design

The study employed a qualitative descriptive design to explore the experiences of family caregivers. This kind of research design is mostly used in investigating a problem that has not been clearly defined (15). It was also important to use this design to obtain information useful in designing others studies. Also, it is important to note that, the design was selected to enhance interaction between the researcher and family caregivers. This interaction helped the researcher to learn from the caregivers’ perspectives.

### Study participants

The participants of this study were family caregivers of patients with HF. In this context, any family member, spouse, friend, or neighbour who had a close personal bond with patients with HF and offered a wide range of support was referred to as a family caregiver. Family caregivers who had been staying with the patient for 14 days and above, able to understand and speak Swahili were included in the study. Family caregivers who were stressed or mentally disturbed during the time of the interview were excluded because they could not provide adequate information as most of the time they thought about their patients.

### Sampling methods and procedure

A purposive sampling method was used to obtain the study participants. In this technique, participants with rich information about caring for their patients were selected to participate. This process suited the purpose of the study as proposed by Etikan et al, (16). The participants were approached during patients’ visiting hours and requested to make an appointment for the data collection at their convenient time and place. The wards in charge, who was conversant with participants’ eligibility criteria were asked to identify potential participants. A sample of 10 participants was considered sufficient because additional participants could lead to the accumulation of unnecessary information (17). The sample size was considered to be sufficient in providing information that helped in meeting the objectives of the study while at the same time ensuring the timely accomplishment of the study.

### Data collection

Data were collected using in-depth interviews. This method provides a deep understanding of a subject under study (18). The data collection took place from April to May 2022. Before data collection, the first author developed an interview guide in response to specific objectives. The guide helped to capture the reality and nature of the research question being studied. The questions were developed from reviewed literature although other questions were raised during the interview as probing questions. The probing questions were prepared to deepen the understanding of the information which caregivers provided. The question in the interview guide includes, what are your needs as a caregiver of a patient with HF? What challenges do you face in caring for your patient with HF? And what support do you get from nurses when it comes to caring for patients with HF? The use of probes helped to get more clarification upon what the participants said and it allowed flexibility and room for extra questions which were not reflected in the guide but still focused on the same topic.

The interview took place at the hospital premises in the small room that the first author booked for the interview. The room was big enough allowing participants free expression and good sitting arrangement. It was also well-ventilated and had good lighting to allow us to observe nonverbal expressions. Furthermore, the room is situated away from the wards and OPD and thereby providing a quiet and calm environment for the interview process. The first author conducted all the interviews. Data were collected using a digital voice recorder and all participants accepted to be recorded. Data collection continued until information saturation was obtained. The interview questions were modified slightly based on new emerging information from the participants. The duration of the interview was between 30 to 45 minutes, and the interviews were conducted in Swahili language

### Data analysis

The analysis of data started just after the first interview. Before analysis, all the recorded information was transcribed verbatim to keep the original meaning of the information. (19) The first author and the research assistant transcribed the interviews. The transcribed information was also checked for consistency from the audio data. A few omissions of information and typos were corrected accordingly. The authors read and re-read the field notes and transcribed data until they had a firm grasp of the data gathered. To-and-back translation of transcripts from Swahili to English as proposed by Braun and Clarke (20) was done to ensure that no information is lost during the process (21).

The thematic analysis approach was employed during data analysis as guided by Braun and Clarke, 2006. This analytical approach was used because we wanted to find out about views, opinions and experiences from the perspectives of family caregivers (20). In this analysis, the first author coded the data manually. This was chosen because the sample size was small and therefore manageable in the available time. The generation of initial codes involved data reduction by the creation of categories that represented the meanings existing in the data segments. Initial codes were produced through terminologies participants used during the interview and used as a reference point for their experiences during the interview. These codes facilitated the researcher’s ability to locate pieces of data later in the process. Concepts were developed by observing pertinent phenomena in the context of the study’s goals. By incorporating, eliminating, merging, or splitting potential codes, we improved codes.

Searching for themes began when all data were initially coded and collated, and a list of different codes was identified across the data set. We used tables to write and name each code and a brief description on a separate piece of paper and played around with organizing them into themes. Individual themes were compared to ensure that they are mutually exclusive from each other. We reviewed and refined the developed themes. In this aspect, we read all the collated extracts for each theme and considered whether they appear to form a coherent pattern. This involved ensuring the validity of individual themes to the data set, but also assessing whether the candidate theme accurately reflected the meanings in the data set as a whole or not. We then defined and named the themes and subthemes **(Table 1)**.

**Table 1:** Example of the coding process.

| Codes | Subtheme | Themes |
| --- | --- | --- |
| Increased patients' social needs<br>Increased patients' physical dependence<br>Lack of money<br>Caregivers' limited time to do business activities<br>Caregivers' inability to pay hospital bills | Failure to meet caregiving needs | New caring role and lifestyle challenges |
| Immobile patient<br>Unstable patient condition<br>Suffering to lift a heavy patient<br>Staying with a patient all days<br>Attention demand due to disease condition | Caring for the critically ill patient |  |

## Ethical Considerations

The Research Ethics Committee of Muhimbili University of Health and Allied Sciences (MUHAS) approved the study with Ref. No MUHAS-REC-03-2022-1057. The permission to collect the data was obtained from the head of research, training, and consultancy of JKCI with Ref. No. AB123/307/01G/50. A signed, written informed consent was obtained from every participant before enrolling in the study. The significance of the study was explained to participants, who were then requested to give informed consent and engaged willingly in the study. Participants were informed of the right to leave the study at any time whenever they wished to. Additionally, participants were informed that there would be no consequences for choosing to withdraw or decline to participate.

## Findings

### Socio-demographic characteristics of participants

Ten participants were included in this study. Six of the participants were females, while four were males. Those under 40 years old accounted for 5 and the rest were above 40 years old. Four participants were single, five were married, and one was a widow. Participants’ educational levels were as follows: five had primary education, three had secondary education, and two had higher education (college education). Three participants were employed, five were self-employed, and two were unemployed.

### Themes and subthemes

Three themes emerged in this study namely; high demand for caring support, new caring role and lifestyle challenges, and professional support **(Table 1)**

**Table 1:** Themes and sub-themes

| Theme | Sub-theme |
| --- | --- |
| Demands for supportive care | The need for social support |
|  | The need for financial support |
| New caring role and lifestyle challenges | Learning to provide the required care at the right time |
|  | Failing to participate in social events |
|  | Caring in an unfavourable environment |
| Professional support | Compliance with professional instruction |
|  | Effective nurse-caregiver interaction |

### Demand for supportive care

Family caregivers reported that caring for patients with HF require their time and financial stability as patient demand a lot of things. This theme is explained under two sub-themes; the need for social support and the need for financial support.

#### The need for social support

Participants reported requiring help to ease their caring responsibilities. They expressed that they need assistance from friends and family members to facilitate a caring role. They also added that feeling lonely while caring for their patients was one of the obstacles to providing effective care to their patients as stated below by one of the participants:

*“I need someone to help me to take care of the patient. For now, there is no one to help me in this task. I wish there was someone to help me”* (Male participant).

Also, participants explained that they needed someone to help them to get capital so that they could do small businesses to sustain their lives and caring responsibilities. One of the participants said:

*“For now I need capital so that I can do small businesses to sustain my life. I can do the light jobs like cooking samosa and mandazi, at least I can improve my life*.*”* (Female participant).

#### The need for financial support

A participant expressed facing financial difficulties that make them fail to meet the daily needs that facilitate caring roles. Financial difficulties were reported as a challenge as they needed money to cover various bills and life needs. One of the participants revealed that:

*“For me, I’ll say the challenge I get is money issues! If you don’t have the money you won’t be able to pay for a bed, when you get to the hospital, you pay there. I came here a month ago, and*

*I stayed for many days, I got the bill, and I knew it was for the bed and the medical costs, I told him I don’t have the money*.*”* (Female participant).

Again, participants expressed that a lack of money was an obstacle in taking patients to regular clinics for checkups and they sometimes had to postpone until they could get money, which could take time. They did not have any money to pay for transport to the hospital as one of the participants described:

“*The challenge is that it can reach the day of clinic and you don’t have any money in your pockets, and the patient is so sick that she cannot walk on foot, so you have to delay going to the clinic*.*”* (Female participant).

### New caring role and lifestyle challenges

New caring roles emerged because their patients needed continuous support. In this case, the participants had to re-adjust their lifestyle and responsibilities of daily living. This posed challenges in fulfilling the two roles and responsivities at the same time. Hence, failing to provide the required care to their patients at the right time. They also expressed failing to participate in social events and caring in an unfavourable environment

#### Learning to provide the required care at the right time

Participants asserted that their patients were dependent and that they required assistance to meet their daily needs. However, they expressed that the caring tasks were difficult when the patient becomes obese making it a challenge to manage the patient alone. One participant said:

*“A patient with a heart condition sometimes requires you to lift him so that he can move or do something, and my patient has become heavy making it difficult for me to lift him alone, and I am worried my health could be at risk too”* (Female participant).

Also, participants stated that patients with HF require ongoing care from them (caregivers). They added that patients required constant attention because their conditions could change at any time and become worse. This made them (caregivers) worried that they could lose their beloved ones at any time. One of the participants reported that:

*“The patient may experience difficulty in breathing, sometimes feel palpitation. This condition disturbs me, and you know I don’t have equipment that can help the patient to breathe*.*”* (Female participant)

#### Failing to participate in social events

Participants described that participation and engagement in their community activities had decreased. They reported that their relationships with their communities have been strained as a result of their responsibilities to their patients. They determined that they do not have time to leave their patients alone. One of the participants stated:

*“I don’t get time to participate. We had a patient at certain hospital, he died and was buried in my region but I could not attend the burrial because of my patient’s condition. He is a man you cannot leave for a minute; I am with him all the time*.*”* (Male participant).

Also, participants reported that due to their caring role it was difficult for them to attend social events such as ceremony because their patient needed them all the time. One of the participants said:

*“It is not easy for sure. I cannot leave my patient. That I should leave the hospital to attend a wedding ceremony? I cannot go anywhere; I am just here with my patient”* (Female participant).

#### Caring in an unfavourable environment

Participants expressed being subjected to a risky and infectious environment when caring for their patients. They explained that there was no toilet specific for caregivers that necessitated them to use the same toilet with the patients. From their perspectives, this phenomenon was putting them at risk of contracting various diseases as stated by the participant below:

*“That is, we [caregivers] who are here are very vulnerable to our health because even the toilets for caregivers are not there. I recently felt my body was weak and every body part was aching*.*”* (Male participant).

Aside from the unfavourable environment in the hospital toilets, participants noted that there was no official place for caregivers to sleep at night. This caused them to sleep while seated on chairs or the floor, or not sleep at all during the night. This made them feel ill because they felt exhausted and experienced generalized body pains. One participant elaborated:

*“At the hospital, there is no official place to sleep, as caregivers, we find it difficult to sleep, and we sit on chairs like this and take a nap. There is no place for you to say you would lie down for a while*.*”* (Male participant).

### Professional support

In the caring role, the family caregiver needs support from a health care professional as handling patients with HF needs skills and knowledge on what to do to improve the patient quality of life. Caregivers reported that compliance with professional instruction and effective nurse-caregiver interaction is an outcomes of good support from professionals.

#### Compliance with professional instruction

Participants stated that information provided by nurses and other health care providers in the hospital was useful for their patients and for them to improve the care that they delivered. They explained that they were told how to feed their patients, give medication to their patients in an emergency before they arrive at the hospital, and how to perform simple physical exercises. One participant expressed that:

*“They tell you the dos and don’ts. If the patient has defecated wear gloves. For example, my patient refused to eat for about five days, they inserted a nasogastric tube into the patient and they directed me on how to feed her, they would feed her three times a day, I learned how to do it but they still kept coming to feed her. For they can get an emergency, I learned and so far I can feed her myself*.*”* (Female participant)

The participants also stated that they had received medical advice from nurses regarding dietary and lifestyle modifications, maintaining follow-up appointments, and completing prescription treatments as directed. This was important for the betterment of the patients as in the following participant’s expression:

*“Instructions you can probably be told to insist on him what the patient should eat, monitor the food he is assigned to eat, treatment and medication compliance. That is the instructions we are given, including how to exercise*.*”* (Female participant).

Moreover, participants appreciated the information provided by nurses and stated that the information not only was beneficial to the patients but also to them (caregivers). One participant revealed that:

*“They come telling you this patient should not be given something; this patient is not this and that. Therefore, they give you a lot of things that help you as a caregiver, and I’m directed to use medication that helps my patient*.*”* (Female participant)

#### Effective nurse-caregiver interaction

Participants reported enjoying the qualities of nursing care at the JKCI. They mentioned that nurses were so collaborative that they responded quickly whenever they were called for help which gave them hope and confidence whenever they needed help. One participant expressed that:

*“I have stayed at the hospital for long time. I am grateful for this hospital; they have given me a lot of support. The patient was in a bad condition, but now he is at least okay. Without them, we could have not reached this day*.*”* (Female participant)

Also, nurses came together and formed a team to solve patient care and other health-related issues to improve patient care quality. Nurses taught caregivers psychology to improve patient health outcomes through good communication and daily relationship building. At the same time, participants appreciated the collaboration from nurses and one of the participants expressed that: *“You tell them this, they listen to you and take care of your patient well; you wish they could continue the service. And we keep in touch. They don’t help me economically, but emotionally and psychologically it helps because you have confidence what my patient is given, gets cured*.*”* (Female participant).

## Discussion

This study explored the experience of family caregivers for patients with HF. It is revealed that caregivers needed social and financial support to facilitate the caring process. Learning to provide the required care at the right time was the new role acquired by caregivers. Failing to participate in social events and caregiving in an unfavourable environment were reported as challenges in caregiving. However, compliance with instruction and effective interaction among the nurses and caregivers were considered to be positive professional support.

Support from family and friends in caring for patients with heart failure is a common social phenomenon as it brings relief to the caregivers and a feeling of well-being(11). In our findings, the lack of social and financial support from family members and friends indicates that the support system in our family is decreasing. This can be viewed in terms of the transition from extended family to nuclear family whereby little or no social support is provided (22) These findings are consistent with the findings of Chi et al, who found it difficult to balance caregiving with other responsibilities and find time to manage their quality of life (23). Also, the findings in this study are consistent with the study by Dionne-Odom et al who reported that caregivers had low familial social support (24). However, there are other factors such as relationship patterns among caregivers, patients and other family members as well as poverty among family members that need further exploration to arrive at a solid conclusion.

Caregivers of HF patients who participated in this study needed help from friends and families in their caregiving role. Caring for patients with HF was a difficult task that needs devotion to undertake and most of the time the ones who are passionate to help, get involved in caring for patients with HF. Caregivers of patients with HF needed to engage in various social activities but couldn’t find the time because they spent most of their time with their patients. This discovery supports earlier findings in Nigeria and Ghana, where the caregiving role affected caregivers’ social engagement (25,26). Doherty et al and Wingham et al reported similar findings (27,28). According to this study, caregivers of HF patients demand financial stability. They are required to pay hospital bills and buy medicines prescribed to their patients. They failed to do so and this caused inadequate patient care. According to this study, a lack of money caused clinic visits to be delayed or even missed entirely. The findings in this study relate to a study which was done in China that reported that lack of money contributed to inadequate care for patients with HF (29). Likewise, a study by Grants and Graven in England revealed a lack of financial well-being was among the challenges faced by caregivers (30). And a study in Zambia by Musonda, et al reported a lack of supporting resources being among the challenges faced by caregivers (31). The result is similar as both studies used a non-informal caregiver of a patient with HF and data were collected using a cross-sectional study design and qualitative study design where data was obtained once so it was easy for family caregivers to verbalize their challenges. Therefore, there is a need for providing adequate support to the family caregiver by identifying and evaluating their needs so that appropriate intervention could be designed to support caregivers and consider an urgent priority.

Also in this study, participants reported learning to provide required care at the right time. An HF patient’s condition can change anytime and a patient may experience difficulty in breathing and palpitations. In this case, a family caregiver may not know what to do to save the lives of their beloved ones. This made caregivers not leave as their patients’ condition could change at any time and their assistance could be of great help. These findings are consistent with a study by Kim and colleagues in Korea which reported similar findings (10). Again, caregivers reported that it was difficult to estimate how long the situation could last, fearing that patient could die. These findings are similar to the study by Tang in China who reported that caring for somebody with a chronic illness makes caregivers worry and fear that the patient is near death and could die at any time (32).

Furthermore, in this study, an unfavourable caring environment while patients are in the hospital has been reported. This may be explained by the fact that social services in the hospital are not adequate. The lack of a formal place for caregivers to rest and sleep at night while they are in the hospital was a distressing phenomenon. Although sharing toilets with patients was considered to jeopardize their health status, there are no other means to extend these services for caregivers. Other studies have reported similar findings. For example, Chukwu and colleagues’ study in Nigeria found infrastructure and amenity deficiencies (25), while in Uganda, inadequate accommodation, sanitation, and the lack of running water were reported as challenges faced by caregivers at the hospital (33).

In this study, support to the family caregivers of patients with HF was important as caregivers reported to have compliance with professional instruction which helped them in caring for their patients with HF. The findings in this study are consistent with the study in the United Kingdom that reported support to caregivers was provided with useful information from nurses. (28). However, the findings of our study differ from studies done in Nigeria and Iran where caregivers reported receiving inadequate support and little guidance from healthcare workers in their caregiving roles (9,25). Another study done in Norway reported that caregivers of patients with HF received insufficient information at the time of discharge from healthcare providers which made caregivers and their patients live unhealthy life and led to multiple readmission (11). The findings in this study might be used to develop strategies and policies which can be used to address the challenges caregivers face when caring for their patients including providing nurses with education which will enhance them to provide sufficient information to caregivers to improve the well-being of the patient and their caregivers.

## Limitations

Caregivers interviewed were those who only managed to come to the hospital for treatment, and those who were home researchers did not manage to explore their experience, so the experience was not broadly explored, therefore, further studies have to be carried out to explore the experience of family caregivers especially those who are at home.

There was a chance that the participant’s original meaning would not be preserved in the Swahili-to-English translation of the transcript. The researcher employed language specialists who are conversant in both Swahili and English to lessen this prejudice. The transcripts of the audiotapes were translated into English and compared to the Swahili transcript for verification.

## Conclusion

The goal of this study was to explore the experiences of the family caregivers of HF patients. Caregivers need social and financial support to provide effective care to their patients. Caregiving is a learning process that needs continuous educational support to adapt to the new caring roles and challenges. Nurses should conduct regular assessments to explore caregivers’ needs, challenges, and concerns and provide timely counselling that can facilitate coping.

## Data Availability

All relevant data are within the manuscript and its Supporting Information files

## Acknowledgement

Special thanks to my study participants; I am delighted for their cooperation, and hospitality, and for providing valuable information on their experiences in caring for patients with HF during my research., Nurse in charge in each unit at JKCI for helping in process of getting the correct participants who suit the purpose of study, and my institution; Jakaya Kikwete Cardiac institute for their support, May God bless you abundantly

## References

1. Bozkurt B, Coats AJS, Tsutsui H, Abdelhamid CM, Adamopoulos S, Albert N, et al. Universal definition and classification of heart failure: a report of the Heart Failure Society of America, Heart Failure Association of the European Society of Cardiology, Japanese Heart Failure Society and Writing Committee of the Universal Definition o. Eur J Heart Fail. 2021;23(3):352–80.

2. Lippi G, Sanchis-Gomar F. Global epidemiology and future trends of heart failure. AME Med J. 2020;5(Ci):15–15.

3. Savarese G, Lund LH. Epidemiology Global Public Health Burden of Heart Failure. 2017;7–11.

4. Kitko L, McIlvennan CK, Bidwell JT, Dionne-Odom JN, Dunlay SM, Lewis LM, et al. Family Caregiving for Individuals with Heart Failure: A Scientific Statement from the American Heart Association. Circulation. 2020;141(22):e864–78.

5. Connolly M, Beattie J, Walker D, Dancy M. End of life care in heart failure A framework for implementation. NHC Improving Quality; 2016. p. 1–28.

6. Agbor VN, Essouma M, Ntusi NAB, Nyaga UF, Bigna JJ, Noubiap JJ. Heart failure in sub-Saharan Africa: A contemporaneous systematic review and meta-analysis. Int J Cardiol [Internet]. 2018;257:207–15. Available from: https://doi.org/10.1016/j.ijcard.2017.12.048

7. Gallagher J, Mcdonald K, Ledwidge M, Watson CJ. Clinical Syndromes Heart Failure in Sub-Saharan Africa. 2018;21–4.

8. Makubi A, Hage C, Lwakatare J, Kisenge P, Makani J. Europe PMC Funders Group Contemporary aetiology, clinical characteristics and prognosis of adults with heart failure observed in a tertiary hospital in Tanzania□: the prospective Tanzania Heart Failure (TaHeF) study. Heart. 2017;100(16):1235–41.

9. Etemadifar S, Bahrami M, Shahriari M, Farsani AK. Family caregivers’ experiences of caring for patients with heart failure: A descriptive, exploratory qualitative study. J Nurs Res. 2015;23(2):153–61.

10. Kim EY, Oh S, Son Y. Caring experiences of family caregivers of patients with heart failure□: A meta-ethnographic review of the past 10 years. Eur J Cardiovasc Nurs. 2020;1–13.

11. Kirsti A, Strom A, Korneliussen K, Fagermoen MS. Family caregivers to a patient with chronic heart failure living at home: co-workers in a blurred hearth care system. Nor J Nurs. 2016;11(2):158–65.

12. Hertz JT, Sakita FM, Kweka GL, Loring Z, Thielman NM, Temu G, et al. Healthcare-seeking behaviour, barriers to care and predictors of symptom improvement among patients with cardiovascular disease in northern Tanzania. Int Health. 2019;1–8.

13. Cleland JGF. Improving care for patients with acute heart failure□: before, during and after hospitalization. 2015;(January):110–45.

14. JKCI. JKCI HOSPITAL OVERVIEW [Internet]. 2020 [cited 2021 Jul 23]. Available from: www.jkci.or.tz/about/category/hospital-overview

15. Boru T. CHAPTER FIVE RESEARCH DESIGN AND METHODOLOGY 5. 1. Introduction Citation□: Lelissa TB (2018); Research Methodology□; University of South Africa, PHD Thesis. 2018;(December).

16. Al, Etikan Ilker, Sulaiman a. Musa RSA. Comparison of Convenience Sampling and Purposive Sampling Comparison of Convenience Sampling and Purposive Sampling. Am J Theor Appl Stat. 2017;11(February).

17. Sim J, Saunders B, Waterfield J, Kingstone T. Can sample size in qualitative research be determined a priori□? Int J Soc Res Methodol [Internet]. 2018;5579:1–16. Available from: https://doi.org/10.1080/13645579.2018.1454643

18. Showkat N, Parveen H. In-depth Interview Quadrant-I (e-Text). 2017;(July).

19. John Creswell JDC. No Title. Fifth edit. Helen Salmon, Chelsea Neve MH, editor. Los Angels; 2018.

20. Braun V, Clarke V. Using thematic analysis in psychology. Qual Res Psychol. 2006;3:77–101.

21. Ho S, Holloway A, Stenhouse R. Analytic Methods ‘Considerations for the Translation of Sensitive Qualitative Data From Mandarin Into English. Int J Qual Methods. 2019;18:1–6.

22. Lloyd-Jones DM, Larson MG, Leip EP, Beiser A, D’Agostino RB, Kannel WB, et al. Lifetime risk for developing congestive heart failure: The Framingham Heart Study. Am Hear Assoc. 2002;106(24):3068–72.

23. Chi NC, Demiris G, Pike KC, Washington K, Parker Oliver D. Exploring the Challenges that Family Caregivers Faced When Caring for Hospice Patients with Heart Failure. J Soc Work End-of-Life Palliat Care [Internet]. 2018; Available from: https://doi.org/10.1080/15524256.2018.1461168

24. Dionne-Odom JN, Hooker SA, Bekelman D, Mcghan G, Kitko L, Wells R, et al. Family Caregiving for Persons with Heart Failure at the Intersection of Heart Failure and Palliative Care: A State-of-the-Science Review. Hear Fail Rev. 2018;22(5):543–57.

25. Chukwu Ngozi, Agwu Prince, Ajibo Henry AN. Challenges faced by informal caregivers of patients in a Nigerian hospital and implications for social work. J Soc Work. 2022;1–18.

26. Agyemang-duah W, Mensah CM, Peprah P, Arthur F, Addai B, Abalo EM. Informal health care□: examining the role of women and challenges faced as caregivers in rural and urban settings in Ghana. J Public Heal. 2018;(2003):1–7.

27. Doherty LC, Fitzsimons D, McIlfatrick SJ. Carers’ needs in advanced heart failure: A systematic narrative review. Eur J Cardiovasc Nurs. 2016;15(4):203–12.

28. Wingham J, Frost J, Britten N, Jolly K, Greaves C, Abraham C, et al. Needs of caregivers in heart failure management: A qualitative study. Chronic Illn. 2015;11(4):304–19.

29. Hu X, Dolansky MA, Hu X, Zhang F, Qu M. Factors associated with the caregiver burden among family caregivers of patients with heart failure in southwest China. Nurs Heal Sci. 2016;18(1):105–12.

30. Grant JS, Graven LJ. Problems experienced by informal caregivers of individuals with heart failure: An integrative review. Int J Nurs Stud [Internet]. 2018;80:41–66. Available from: http://dx.doi.org/10.1016/j.ijnurstu.2017.12.016

31. Musonda KC, Nyashanu M, Mutale W, Sitali D, Mweemba O. Exploring The Challenges Faced by Informal Home Based Palliative (HBP) Caregivers in Ndola District, Exploring The Challenges Faced by Informal Home. J Soc Work End Life Palliat Care [Internet]. 2021;17(4):349–63. Available from: https://doi.org/10.1080/15524256.2021.1976351

32. Tang Y. Caregiver burden and bereavement among family caregivers who lost terminally ill cancer patients. Paliative Support Care. 2018;1–8.

33. Sadigh M, Nawagi F, Sadigh M. The Economic and Social Impact of Informal Caregivers at Mulago National Referral Hospital,. Ann Glob Heal [Internet]. 2016;82(5):866–74. Available from: http://dx.doi.org/10.1016/j.aogh.2016.06.005

